# German Recommendations for Physical Activity and Physical Activity Promotion in Adults with Noncommunicable Diseases

**DOI:** 10.1101/19008953

**Authors:** Wolfgang Geidl, Karim Abu-Omar, Mayra Weege, Sven Messing, Klaus Pfeifer

**Affiliations:** Department of Sport Science and Sport, Division of Exercise and Health, Friedrich-Alexander University Erlangen-Nürnberg, Germany; Department of Sport Science and Sport, Division of Physical Activity and Public Health, Friedrich-Alexander University Erlangen-Nürnberg, Germany; Committee for the Development of the German Recommendations on Physical Activity and Physical Activity Promotion

**Author notes:** **Corresponding author:** Dr Wolfgang Geidl, Department of Sport Science and Sport, Friedrich-Alexander University Erlangen-Nürnberg, Gebbertstraße 123b, 91058 Erlangen, Germany.

**Keywords:** Guidelines, Recommendations, Physical Activity, Physical Activity Promotion, Behaviour Change, Noncommunicable Disease, Chronic Condition

## Abstract

**Background:** The objective of this study was to develop evidence-based recommendations for physical activity (PA) and PA promotion for German adults (18–65 years) with noncommunicable diseases (NCDs).

**Methods:** The PA recommendations were developed based on existing PA recommendations and using a three-phased process. In phase 1, systematic literature searches were conducted for current PA recommendations for seven chronic conditions (osteoarthrosis of the hip and knee, chronic obstructive pulmonary disease, stable ischemic heart disease, stroke, clinical depression, and chronic non-specific back pain). In phase 2, the PA recommendations were evaluated on the basis of 28 quality criteria. High-quality recommendations were identified, and a content analysis was conducted on these recommendations. In phase 3, the findings of the content analysis were summarised, and PA recommendations for seven chronic conditions were deducted. The seven recommendations were then synthesised to generate generic German PA recommendations for adults with noncommunicable diseases (NCDs). In relation to the recommendations for PA promotion, a systematic literature review was conducted on papers that reviewed the efficacy/effectiveness of interventions for PA promotion in adults with NCDs.

**Results:** The German Recommendations for Physical Activity state that adults with NCDs should, over the course of a week, should do at least 150 minutes of moderate-intensity aerobic PA, or 75 minutes of vigorous-intensity aerobic PA, or a combination of both. Furthermore, muscle-strengthening activities should be performed at least twice a week. The promotion of PA among adults with NCDs should be theory-based, specifically target PA behaviour, and be tailored to the respective target group. In this context, and as an intervention method, exercise referral schemes are one of the more promising methods of promoting PA in adults with NCDs.

**Conclusion:** The development of evidence-based recommendations for PA and PA promotion is an important step in terms of the initiation and implementation of actions for PA-related health promotion in Germany. The German Recommendations for PA and PA promotion inform adults affected by NCDs and health professionals on how much PA would be optimal for adults with NCDs. Additionally, the recommendations provide professionals entrusted in PA promotion the best strategies and interventions to raise low PA levels in adults with NCDs. The formulation of specific PA recommendations for adults with NCDs and their combination with recommendations on PA promotion is a unique characteristic of the German recommendations.

## INTRODUCTION

The high prevalence of physical inactivity is a global problem [1, 2] that contributes to increasing morbidity, higher rates of premature death [3], and increased economic costs [4]. In this context, the development of strategies to promote physical activity (PA) is an important challenge, both globally and nationally. A method of combatting high levels of inactivity is the development of PA guidelines. PA guidelines define the amount of health-enhancing activity through which significant health gains can be achieved [e.g. 5]. Both the World Health Organization (WHO) and the European Union have urged their member states to develop their own recommendations on a national level [5–8]. Even though many nations have developed these recommendations in recent years, including Canada, Great Britain, Australia, Switzerland, and Australia [e.g. 9, 10], the resulting guidelines predominantly focus on age-stratified target groups of children and adolescent, adults, and older adults. Thus far, the target group of individuals with noncommunicable diseases (NCDs) has received scant consideration.

The fact that there are hardly any national PA recommendations for adults with NCDs is surprising and problematic for several reasons. The ever-growing prevalence of NCDs [11] has become a global health issue. For most countries, adults with NCDs comprise one of the largest population groups. In Germany, for example, four out of every 10 adults report themselves as having at least one NCD [12]. PA has been proven to not only aid in the prevention [13] but also in the treatment of NCDs [14]. For more than 25 NCDs – such as type 2 diabetes mellitus, osteoporosis, ischaemic heart disease, and clinical depression – PA is viewed as a medicine [15]; a medicine that positively influences symptoms and comorbidities, physical fitness and health-related quality of life [15]. Therefore, WHO [16] states that regular PA is the ‘best buy’ in controlling NCDs. Accordingly, WHO [5] specifies that their PA recommendations that are relevant for all healthy adults ‘also apply to adults with chronic noncommunicable conditions not related to mobility such as hypertension or diabetes.’ The PA recommendations of WHO are based on a vast body of evidence, and they have an immensely positive influence on global actions regarding PA promotion. However, a limitation of WHO’s recommendations is that their underlying scientific evidence is mainly based on the general population, meaning that specific scientific evidence regarding the health effects of PA on adults with NCDs has not been systematically considered. This is problem if adults with NCDs would need other PA and different levels of PA to optimally promote their health. This lack of disease-specific evidence in PA recommendations also applies to nations that base their PA recommendations for adults with NCDs on WHO recommendations (e.g. Austria, Ireland, and Sweden). Other countries in Europe (e.g. Netherlands) excluded studies in people with NCDs in developing their national PA recommendations but state that the recommendations are useful for numerous specific groups of people with a chronic condition [17]. And some countries (e.g. Belgium, Greece, Spain, and Great Britain) do not make specific exercise recommendations for adults with NCDs [11]. This means that for many countries, there exists no evidence-based health-promoting PA recommendations for the large group of adults with NCDs. Thus, neither the adults affected by NCDs nor the health professionals involved in PA promotion know how much PA would be optimal.

Even more uncommon than missing PA recommendations, however, is to identify guidelines which not only define the health-enhancing dose of PA, but also recommend how PA promotion works for the specific target groups of adults with NCDs [18]. The majority of countries fail to provide such recommendations on PA promotion for adults with NCDs within a specific national scenario. Therefore, health professionals entrusted in PA promotion often lack recommendations for the best strategies and interventions to raise low PA levels in adults with NCDs.

With such limitations in mind, Germany developed its own national recommendations in 2016 [19]. The Committee for the Development of the German Recommendations on Physical Activity and Physical Activity Promotion consisted of an interdisciplinary working group made up of 16 scientists from six German universities. From an international perspective, the German recommendations are characterised by two unique features – namely, the formulation of specific recommendations for adults with NCDs and the systematic integration of guidelines for PA and PA promotion for adults with NCDs as well as for the other target groups (children and adolescents, adults, and older adults).

Following on from this, the purpose of this paper is to describe the methodology we used to develop the German National Recommendations for Physical Activity and Physical Activity Promotion in Adults with NCDs and summarise the main results of this development. By doing this, the paper hopes to help other nations develop their own PA guidelines for the increasingly relevant target group of adults with NCDs. The paper also hopes to promote debate in relation to the implications of addressing specific target groups and the impact of this specific targeting on PA levels and public health policies.

## METHODS

### Physical activity recommendations

For the development of the German PA recommendations for all target groups (children and adolescents, adults, older adults, and adults with NCDs), the same three-phase process was used (see Table 1).

**Table 1.** Methodology used to prepare the German Recommendations for Physical Activity.

|  |  |
| --- | --- |
| **Phase 1** | **1A:** Systematic literature review of existing PA |
| January–June 2015 | recommendations |
|  | **1B:** Survey of experts to establish quality criteria |
| **Phase 2** | **2A:** Assessment of the identified PA recommendations using |
| May–August 2015 | quality criteria |
|  | **2B:** Selection of high-quality PA recommendations as source |
|  | recommendations |
|  | **2C:** Content analysis of the source recommendations |
| **Phase 3** | **3A:** Synthesis of content analyses and derivation of the |
| September–December | recommendations for health-effective PA |
| 2015 |  |

For adults with NCDs, the three-phase process was initially applied to PA recommendations that were specific to the seven NCDs. Considering years of life lost due to premature death as well as years lived with disability, six of these NCDs are among the most burdensome diseases in Germany for both men and women [20]. The six NCDs are clinically stable ischemic heart disease, chronic non-specific back pain, clinical depression, type 2 diabetes mellitus, chronic obstructive pulmonary disease(COPD), and stroke (> 6 months after the acute event). Additionally, osteoarthritis was added as seventh disease. Along with chronic non-specific back pain, osteoarthrosis accounts for the majority of diseases in the disease group of musculoskeletal diseases which is among the three most relevant disease groups regarding burden of disease [20]. Osteoarthritis is characterised by high prevalence and high levels of stress among those affected. Its negative effects include pain, functional restrictions in everyday life, and loss of quality of life [21]. Finally, generic PA recommendations for adults with NCDs were synthesised from an aggregation of the seven disease-specific PA recommendations. We previously published a comprehensive description of the entire methodology [22, 23]. The central aspects of the methodological approach are described below.

#### Phase 1: Systematic literature review and establishment of quality criteria

Separate literature searches were carried out for the seven NCDs (1A). The searches were comprised of PA recommendations, reviews of PA recommendations, and meta-analyses of the effects of PA, all published between 2010 and 2015 on the Medline database, either in German or English. Primary studies were not included.

As a first step in the selection of papers, titles and abstracts were checked for relevance. At this point, primary studies, non-human articles, papers not available in German or English, and papers that dealt with a different topic were excluded. Those papers deemed relevant were then analysed in their entirety. The full texts were subjected to a detailed relevance test that was conducted based on a standardised assessment sheet.

After checking the papers for the correct target population (adults with one of the seven relevant NCDs), all papers containing original PA recommendations for those adults were included. If a paper did not contain any original PA recommendations but contained a meta-analysis of the effects of PA for the relevant target group or a review of clinical guidelines or PA recommendations, then it was included.

For the standardised quality evaluation of the researched PA recommendations, an evaluation instrument was developed (1B). The evaluation instrument was used to ensure that methodically high-quality PA recommendations were selected. First, a list of 23 potential quality criteria was compiled based on the methodology of the *German Instrument for Methodological Guideline Appraisal* [24] and the instrument for *Advancing guideline development, reporting and evaluation in health care* (AGREE) methodology [25]. Second, the list of quality criteria was submitted to national experts for validation, based on the methodological approach of the Delphi surveys [26]. The experts were asked to check the proposed quality criteria for completeness and comprehensibility and to add any missing criteria (Delphi procedure stage 1). Finally, the revised list of quality criteria was submitted to the experts once again to evaluate its relevance with regard to the identification of high-quality PA recommendations (Delphi procedure stage 2).

#### Phase 2: Evaluation, selection, and content analysis of identified physical activity recommendations

The yielded PA recommendations were evaluated based on their content and methodological quality using the evaluation instrument developed in phase 1 (2A). The domains that were evaluated were the scope and purpose of the recommendations, the methodological accuracy in their development, their clarity and differentiation of content, and their structure (see Additional file 1).

The quality evaluation formed the basis for the selection of what is here referred to as the source recommendations (2B). The source recommendations are those used as the basis for the German PA recommendations. The cut-off value for the selection of high-quality recommendations was > 60% in the domains of *scope and purpose* and *methodological accuracy in the development of the recommendations*. Additionally, reviews of recommendations and meta-analyses for the target group of adults with NCDs were included as supplementary resources. We did this without explicit quality ratings because even reviews with low quality could include statement on high quality PA recommendations and meta-analysis normally include a quality rating leading to the inclusion of studies with a high methodological quality. The identified source recommendations were then subjected to a detailed content analysis using a standardised analysis sheet that contained all 28 quality criteria (2C).

#### Phase 3: Synthesis of content analysis and derivation of the recommendations for health-effective physical activity

Based on the detailed content analysis of the source recommendations and supplementary texts, seven disease-specific recommendations for health-relevant PA were compiled. These recommendations were then critically reviewed in relation to the reported health effects, dose-response relationships, and risk–benefit considerations of PA. Finally, the identification of cross-disease commonalities across the seven diseases (e.g., similar amounts of PA or similar types of PA recommended) was discussed resulting in the derivation of generic PA recommendations for adults with NCD.

### Physical activity promotion recommendations

A systematic review of reviews regarding the efficacy/effectiveness of interventions for PA promotion in adults with NCDs was conducted. This review comprises one of the main pillars that led to the development of the German PA promotion recommendations for adults with NCDs.^1^ The methodology used for the systematic review of reviews is briefly described below. A more detailed characterisation has already been explicated [29, 30].

#### Systematic review of review papers that examined the efficacy/effectiveness of interventions for physical activity promotion

A systematic review of review papers that examined the efficacy and effectiveness of interventions was conducted. Six electronic databases (PubMed, Scopus, SPORTDiscus, PsycINFO, ERIC, and IBSS) were systematically searched for the terms ‘physical activity,’ ‘intervention,’ ‘evidence,’ ‘effect,’ ‘health,’ and ‘review.’ Alternative terms were also utilised for PA, including ‘bike,’ ‘biking,’ ‘cycling,’ ‘walking,’ ‘active transport,’ ‘human-powered transport,’ ‘sedentary,’ ‘exercise,’ and ‘sport.’ Two independent reviewers screened titles and abstracts for relevance based on the following criteria: a) the paper contains empirical results from single studies; b) the paper includes interventions focused on PA promotion or the reduction of physical inactivity; c) the paper focuses on the efficacy of interventions; and d) the paper is written in English or German. For the identified relevant papers, a secondary screening process of the entire text was conducted by two independent reviewers based on the same criteria. Finally, additional papers were identified via screening the reference lists of identified articles.

An independent researcher assessed the quality of the identified papers using two separate tools – namely, the AGREE instrument (The AGREE Collaboration 2003), which was used in the formulation of the Canadian Physical Activity Guidelines [31], and our own quality criteria, which allowed for higher levels of differentiation regarding the methodological quality criteria of each review (see Additional file 2). Based on both instruments, percentage values were calculated for each paper. Such values indicated the percentages of fulfilled criteria for each paper, both on the basis of the AGREE tool and our newly developed instrument. These percentage values were calculated based on the number of applicable criteria (e.g. some criteria were only applicable for meta-analyses). The combined results defined the quality of each paper as high, medium, or low. A paper was classified as being of high quality when at least 75% of the AGREE criteria and 60% of our own criteria were fulfilled. Papers were defined as being of medium quality if they satisfied just one of these thresholds, while low-quality papers reached neither threshold.

#### Expert consensus

A group of scientific experts developed the German Recommendations for Physical Activity Promotion in a process of consensus. Two individuals with expertise in PA promotion were assigned the task of assessing the efficacy/effectiveness of the identified papers. Following the methodology proposed by Smith et al. [32], both reviewers applied a standardised process of analysis that consisted of six steps. First, an independent review of the identified literature was conducted, and a draft summary statement was compiled. Second, a meeting of both reviewers was held to discuss statements and agree on a conjointly revised summary statement. Third, the summary statement was presented and discussed with the reviewers who were assigned to other target groups (e.g. older adults). Fourth, a workshop meeting was held to present each summary statement to the entire project group (including scientists involved in drafting the PA promotion recommendations as well as an International Scientific Advisory Board). Each summary statement was revised on the basis of expert feedback. Fifth, the recommendations for each target group were drafted using the finalised summary statements. A template specifying how to draft the recommendations was developed and provided by the project leaders. Sixth, the drafted recommendations were circulated for review by the entire project group as well as the International Scientific Advisory Board. Recommendations were made when both reviewers rated the available evidence as being of strong or medium quality, based on the following criteria: (1) the number of available reviews focusing on a given intervention type is sufficient to formulate recommendations; and (2) the reviews show conclusive evidence for efficacy. Recommendations were not made when the above criteria were not fulfilled (i.e. when the available evidence was weak or inconclusive).

## RESULTS

### Physical activity recommendations

#### Existing physical activity recommendations for adults with NCDs

The PA recommendations for adults with NCDs were derived from a detailed content analysis of 48 source recommendations and texts: arthrosis (hip and knee)(9 articles); type 2 diabetes mellitus (4 articles); COPD (5 articles); clinically stable ischemic heart disease(4 articles); stroke (10 articles); clinical depression (7 articles); and chronic non-specific back pain (9 articles). The detailed references of all 48 articles and their quality ratings can be found in Additional file 3.

For all seven analysed NCDs, PA and/or exercise is recommended as an effective treatment option. The positive benefits of PA outweigh its costs and side effects. A physically inactive lifestyle is associated with significantly greater health risks than a lifestyle with high levels of PA. Details regarding the health effects, risks, and side effects of PA within the individual disease-specific PA recommendations are reported by Rütten et al. [19].

#### Main recommendations for physical activity

The following recommendations are for adults aged between 18 and 65 with an NCD such as type 2 diabetes, COPD, arthritis in the hip or knee, clinically stable ischemic heart disease, stroke (> 6 months after the acute event), clinical depression, or chronic non-specific back pain.

In order to achieve significant health effects (e.g., improved symptoms, enhanced physical functioning, improved psychological health and quality of life), adults with NCDs should be physically active on a regular basis. However, health-enhancing results can also occur when individuals who were entirely physically inactive become somewhat more active. Every step away from physical inactivity is important, as each step leads to increased health benefits.

The PA recommendations for adults with NCDs are presented in Table 2. They do not differ from the PA recommendations for healthy adults and represent the minimum levels of PA one should meet in order to maintain and promote one’s health comprehensiveley.

**Table 2.**
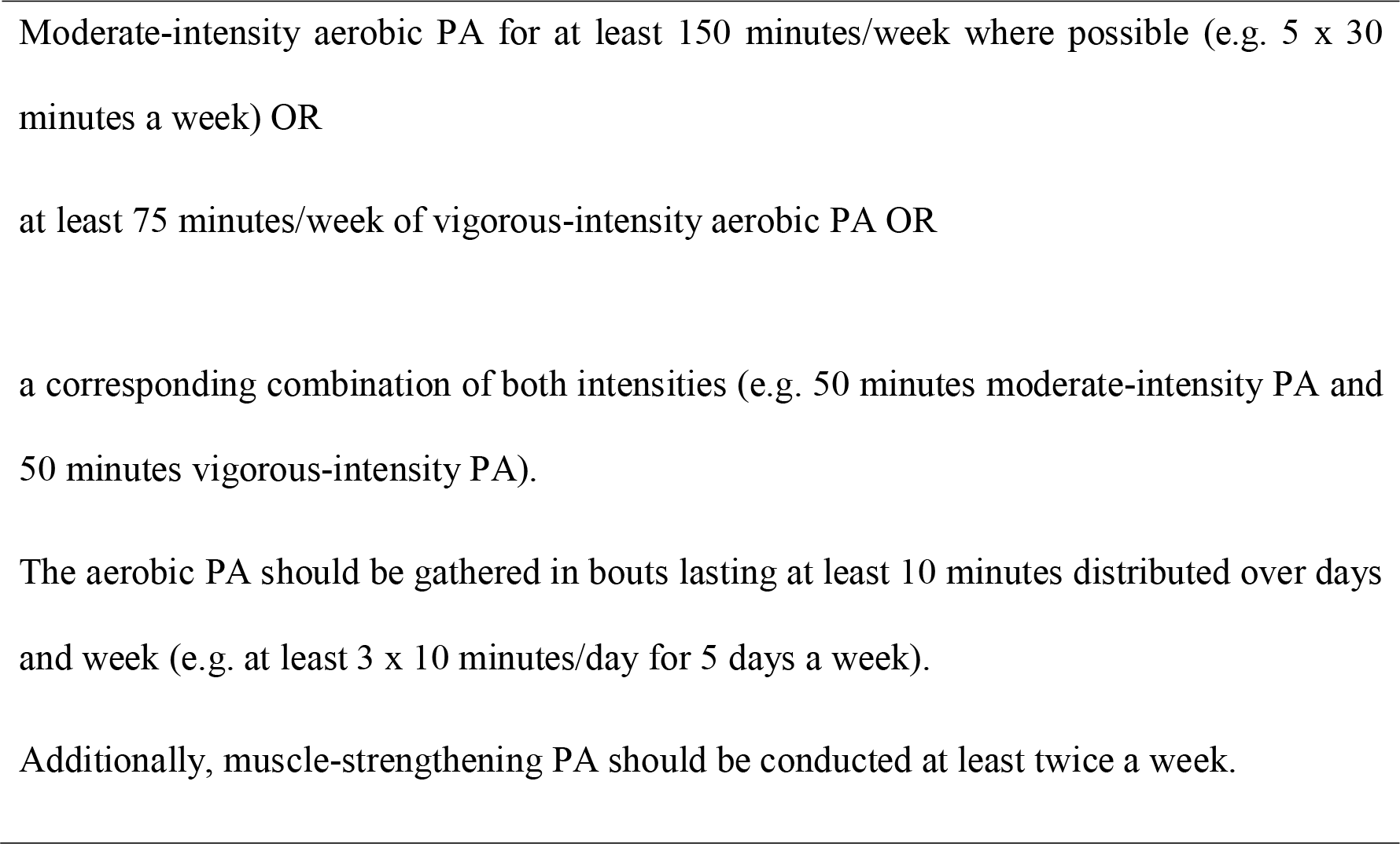
German recommendations for physical activity for adults with noncommunicable diseases.

During phases in which the recommendations cannot be met (e.g. due to severity of illness or reduced physical capacity), adults with NCDs should be as active as their current situation permits.

#### Additional recommendations for safe (re-)entry into a physically active lifestyle

PA is associated with a variety of positive health effects for adults with NCDs. However, PA is not completely without risk for such individuals. The beginning of a physical training programme or phases during which PA levels are increased can be associated with a higher risk of side effects and adverse events. Therefore, the developed recommendations also include information on safety enhancement. To increase the safety and effectiveness of PA, adults with NCDs should:

1. have a (sports) medical examination carried out when commencing a physically active lifestyle or entering a PA programme;
2. decide, along with a doctor, whether practicing PA independently is safe and appropriate, or whether it is advisable to be under the professional care of PA professionals at the outset;
3. tailor the PA dose (type of PA, exercise intensity, duration, frequency) to individual needs and functional levels with the assistance of a PA professional; and
4. obtain professional advice from healthcare professionals during the phases of the progression of the illness, when experiencing lack of control over the illness, or when one’s health status is deteriorating, as it may be necessary to adjust or change PA or momentarily interrupt PA.

### Physical activity promotion recommendations

#### Efficacy/effectiveness for physical activity promotion

The PA promotion recommendations for adults with NCDs were developed based on 18 systematic reviews. The detailed quality ratings of the papers are included in Additional file 4. All the papers conducted their reviews in healthcare settings. Two analysed general PA promotion interventions [33, 34], seven focused on indication-based PA promotion [35–41], three assessed interventions in primary care [42–44], and six dealt with the effects of different types of PA-promoting interventions [45–50].

The reviews analysing general interventions for PA promotion found that the effects of interventions are stronger when PA behaviour is targeted specifically rather than in combination with other health behaviours [33]. Furthermore, interventions are most effective when they are theory-based and use techniques of behavioural change [34].

Six indication-based PA-promotion approaches provided medium-level evidence during the assembling of the recommendations. Short et al. [35] evidenced that special behaviour-based techniques can be useful for adults with breast cancer. Interventions for adults with rheumatoid arthritis were investigated in two reviews [36, 37], and one review indicated that PA promotion for this target group can be effective in the long term [37]. The research findings identified only one study that showed positive effects of a PA-promoting intervention for people with cystic fibrosis [38]. One review analysed home-based exercise programmes for adults with chronic low back pain, and most of the studies reviewed showed significant effects [39]. For adults after stroke, specific behavioural interventions, such as targeted counselling or specially tailored exercise programmes, are more effective than exercise programmes and general counselling [40]. The usual rehabilitation measures comprising PA therapy combined with psychosocial or educational interventions can increase PA behaviour in the short term for individuals with cardiovascular disease [41].

Concerning interventions in primary care and/or curative care, one review shows that exercise referral schemes can lead to small positive effects in the short and medium term [42]. Orrow et al. [44] conducted a review and meta-analysis based on a broad range of interventions and showed that interventions in primary care can be effective. Another review concluded that motivational interventions are effective for PA promotion [43].

The last group of reviews focused on the effectiveness of different types of interventions. Based on these reviews, there is strong evidence for the effectiveness of pedometer-based interventions for adults with musculoskeletal diseases [45], inconclusive evidence for the effectiveness of internet-based interventions in cardiac rehabilitation [46, 47], and conflicting evidence for the effectiveness of web-based interventions for patients with NCDs [50]. Furthermore, the reviews showed evidence of the effectiveness of the counseling method *motivational interviewing* for individuals with NCDs [48] and of lifestyle interventions delivered by nurses in primary healthcare [49].

#### Recommendations for physical activity promotion: efficacy/effectiveness

All papers in the systematic review refer to health care settings, e.g. general practitioners practices or clinics. Accordingly, recommendations for PA promotion in adults with NCDs were made for interventions in healthcare institutions (see Table 3).

**Table 3.** German recommendations for physical activity promotion in adults with noncommunicable diseases.

|  | Healthcare institutions |
| --- | --- |
| Efficacy/effectiveness | Introduce exercise referral schemes |
| and quality criteria | Use theory-based approaches |
|  | Tailor PA behaviour of adults with NCDs specifically |
|  | Tailor interventions to the respective target group |

## DISCUSSION

The German guidelines for PA and PA promotion for adults with NCDs provide recommendations for the amount of PA necessary to yield substantial health benefits and strategies for PA promotion so that the recommended level of PA is successfully reached. Thus far, and to the best of our knowledge, such a combination of recommendations is the first of its kind. The concomitant implementation of both PA and PA promotion recommendations produces considerable health gains in what is, to date, one of the largest and most inactive adult subgroups.

The German recommendations for PA and PA promotion represent a fundamental building block in terms of mobilising national efforts to fight against physical inactivity. On the one hand, professional organisations in the fields of medicine, exercise therapy, and/or rehabilitation can use these guidelines to promote increased PA levels among individuals with NCDs. On the other hand, health professions and societies ought to support the dissemination and implementation of these guidelines in order to maximise their influence and ensure that the target population is reached.

The ideal dosage of PA for various target groups has been long discussed. Evidence supporting the claim that PA promotes health among people with NCDs is plentiful [15]. When considering a wide scope of health outcomes, many adults with NCDs respond to higher levels of PA by producing better health effects, and the understanding that ‘a lot helps a lot’ still rings true. Nonetheless, there are other outcomes in this regard. For example, a higher level of PA intensity might have negative effects on psychological well-being [51]. For some outcomes, much of the maximal effect is achieved from a relatively low dose of PA; for example, the greatest reduction in mortality rates occurs at PA levels already well below the recommended minimum dose of 150 minutes/week [52]. The German PA recommendations for adults with NCDs are based on a synthesis of the available recommendations. Consequently, they also use the concept of the minimum dose for substantial health gains. Nevertheless, it is a task for future researchers to question the concept of minimum doses and determine the dose–response relationships for certain health outcomes (e.g., mortality, morbidity, physical functioning, psychological health and well-being).

The current German PA recommendations for individuals with NCDs are identical to those suggested for healthy adults (i.e. 150 minutes of moderate PA plus two strength-training sessions per week). This generic outcome was synthesised based on seven carefully prepared, high-quality, indication-specific recommendations. The claim that this dose leads to substantial health gains in most individuals with NCDs is evidence-based. Notwithstanding, this dose might be physically and psychologically excessive for some individuals with NCDs – this point cannot be ignored. The importance of patient-centred tailoring plans and programmes that focus on adjusting the dosage to individual needs and functionality levels is therefore emphasised. This supports and values adults with NCDs being regularly active, according to their current health status and their ability.

In addition to PA behaviour, other health-related behaviours, including smoking, alcohol, diet, and medication, have a major influence on the health of individuals with NCDs [53]. It is understood and anticipated that the recommendation to change only one behaviour at a time might not be practical to implement in a real-world setting. For example, in a typical rehabilitation context, it is often recommended to try and adjust multiple behaviours at once. Even if this is medically recommended and meaningful, the probability of successful behaviour change decreases as a result. Influencing multiple behaviour patterns individually might be a better approach. Addressing the promotion of PA first might be the best option, as regular PA can act as a catalyst to promote changes in other health behaviours [54].

The basic premise of the Committee for the German Recommendations for Physical Activity and Physical Activity Promotion was to develop recommendations for specific target groups. Independent and separate working processes were implemented for children and adolescents, adults, older adults, and adults with NCDs. Thus, the German recommendations include more diverse population subgroups and fulfil a requirement of the Advisory Committee of the United States [55]. Due to the rather low level of evidence of PA promotion in individuals with NCDs, the recommendations for adults with NCDs are less detailed compared to the other target groups [29]. Nevertheless, additional to the development of the target-specific recommendations for PA promotion, general quality criteria for the conceptualization, implementation, and evaluation of interventions for PA promotion were developed [27]. For example, during the conceptualisation phase, quality criteria, such as the multidimensionality of the intervention, the involvement of different stakeholders, and the specification of goals and target behaviour, need to be considered. Other criteria relate to, for example, communication, sustainability, and resources during implementation as well as to different aspects of the evaluation (see Additional file 5 for a list of all quality criteria). Even though there is no specific evidence in relation to adults with NCDs for most of these criteria, the criteria support the conceptualisation, implementation, and evaluation of interventions for adults with NCDs.

It is important to highlight that nations should not only *develop* PA and PA promotion recommendations. In order to maximise the reach of the recommendations, it is crucial to attempt to *disseminate* said recommendations to the appropriate individuals, communities, and organisations at the right time. In order to facilitate understanding of the developmental process for PA recommendations, it has been proposed that the German context should be analysed through the lens of the multiple streams approach (MSA) [18]. In summary, the MSA was developed in 1984 to study policy processes and to clarify why certain issues receive or attract attention and others do not (agenda setting). It posits that the political process comprises three streams that flow independently – namely, the problem stream (i.e. specific issues perceived as problematic and in need of solution), the policy stream (i.e. the development of strategies and possible policies to address the stated problem), and the politics stream (i.e. operating actors such as political parties, institutions, and interest groups). The interaction between the three streams might facilitate or hinder the effective dissemination and adoption of national PA and PA promotion guidelines. As for the German context, including PA promotion as a stand-alone topic in the political agenda might have come as a result of said interaction. This propelled several different strategies and actions that boosted the impact of PA and PA promotion recommendations nationwide. A more detailed description and dissection of the aforementioned strategies and actions was published by Rütten et al. [18].

### Limitations

The methodological approach used to develop both guidelines is not without its restrictions. A goal of the Committee for the Development of German Guidelines for Physical Activity was to address the target group of adults with NCDs. However, only a (very) limited amount of finances, time, and human resources was made available for the overall development of the guidelines.

Due to said limitations, the committee opted to apply an overall methodological approach based on the extraction and synthesis of high-quality reviews, meta-analyses, and existing recommendations. Original scientific works, for instance, could unfortunately not be taken into account during this development. The consideration of these works would have allowed for additional and relevant topics to be explored in more detail. As an example of this consequence, the current PA guidelines do not incorporate the role of sedentary behaviour and its impact on adults with NCDs.

As for the PA promotion guidelines, general recommendations can be provided for interventions in terms of their theoretical foundations. However, investigating the most effective theory-based intervention or conducting comparative analyses of the effects of different theories was beyond the scope of our methodological approach.

The underlying evidence comes exclusively from interventions carried out in the healthcare system. Studies carried out in other settings (e.g., educational system, work place) are lacking. It is likely that the consideration of original scientific papers would have identified further studies conducted in other settings. It is likely that this would have led to more multi-faceted recommendations for the promotion of PA in adults with NCDs.

The robustness of the selected methodology as well as its economic limitations represent a compromise that aims to include individuals with different NCDs, as they represent a crucial target group from a public health perspective. When compared to Canada, for example, Germany used a low-cost approach, which resulted in numerous methodological limitations. Such an approach, however, seems to be feasible for other nations, as it helps to curtail restraints imposed by a scarcity of available resources.

## CONCLUSION

The development of PA recommendations for adults with NCDs provides an evidence-based target dose of PA that adults with different NCDs can use to achieve significant health outcomes; the development of PA promotion recommendations for adults with NCDs helps to achieve that dose. Thus, the development of evidence-based PA and PA promotion guidelines is an important building block in relation to the initiation and implementation of actions for PA-related health promotion in Germany. The formulation of specific PA recommendations for adults with NCDs and their combination with specific PA promotion recommendations renders the German recommendations quite unique. The German Recommendations for PA and PA promotion for adults with NCDs help other nations develop their own PA guidelines for the increasingly relevant target group of adults with NCDs.

## Data Availability

Not applicable

## List of abbreviations

COPD: chronic obstructive pulmonary disease
MSA: multiple streams approach
NCD: noncommunicable disease
PA: physical activity
WHO: World Health Organization

## DECLARATIONS

### Ethics approval and consent to participate

Not applicable

### Consent for publication

Not applicable

### Availability of data and material

Not applicable

### Authors’ contributions

This work summarises aspects of the development of the National Recommendations for Physical Activity and Physical Activity Promotion. WG, KA, SM, and KP are members of the Committee for the Development of the German Recommendations on Physical Activity and Physical Activity Promotion. WG and KA conceptualised this paper. WG and KP are responsible for the methods and results related to the German Recommendations for Physical Activity; KA, MW, SM, and KP are responsible for the methods and results related to the German Recommendations for Physical Activity Promotion; MW, KA, and WG wrote the draft of this manuscript; all authors revised the draft of the manuscript and approved the submitted version.

## Acknowledgements

We would like to thank all members of the Bewegungsförderung im Alltag (Physical Activity Promotion in Daily Living) working group who made a valuable contribution to the recommendations through their constructive suggestions and feedback.

## Additional files

Additional file 1.pdf; Criteria for Quality Assessment and Content Analysis of Physical Activity Recommendations. This file lists the 28 criteria used to assess methodological quality and analyse motion-related content.

Additional file 2.pdf; Quality checklist for the systematic reviews of reviews regarding the effectiveness/efficacy of PA promoting interventions. This file lists the criteria used to evaluate the papers reporting PA promotion interventions with regard to their effectiveness/efficacy.

Additional file 3.pdf; Quality ratings of the physical activity recommendations and additional ressources (meta-analysis and reviews on physical activity recommendations) that were additional included without explicit quality ratings for seven noncommunicable diseases. This file lists the retrieved articles and their quality ratings leading to the decision on which articles the German Recommendations for Physical Activity for Adults with NCD were developed.

Additional file 4.pdf; Quality rating of the reviews for developing the German Recommendations for PA Promotion. This table contains the quality rating of the reviews used to develop the German Recommendations for PA promotion.

Additional file 5.pdf; Quality criteria for the conceptualization, implementation and evaluation of PA promoting interventions. This table lists the general quality criteria for PA promotion interventions.

## Footnotes

1 Additionally, two reviews were carried out to analyse the cost-effectiveness of PA promotion interventions and develop generic quality criteria for the conception, implementation, and evaluation of interventions for the promotion of PA. The results of these reviews have been published [27, 28] and are beyond the scope of this article.

